# High prevalence of varicella zoster virus infection among persons with suspect mpox cases during a mpox outbreak in Kenya, 2024

**DOI:** 10.1101/2025.04.08.25325502

**Authors:** Clayton Onyango, Jonas Z. Hines, Melvin Ochieng, Derrick Amon, Shirley Lidechi, Elekiah Anguko, Bonventure Juma, Jeremiah Nyaundi, Morine Achieng, Abdi Roba, Abubakar Hussein, Ahmed Abade, Pius Mutuku, Lucy Munyeki, Fredrick Mutisya, Barbara Jepkorir, Emmanuel Okunga, Daniel Lang’at, John Kiiru, Kimberley D. McCarthy, Amy Herman-Roloff, Naomi Lucchi

## Abstract

During a mpox outbreak in Kenya, 61% of patients suspected of infection tested positive for varicella zoster virus, which causes a rash similar to the mpox rash. Expanding laboratory capacity during the outbreak enhanced disease intelligence by confirming another etiology of rash among patients that tested mpox negative.

---

Since July 2024, Kenya has been experiencing a clade Ib mpox outbreak, with 28 confirmed cases reported through December 11, 2024. To support its public health response, the Ministry of Health (MOH) developed case definitions to guide the testing of persons with suspected mpox.^1^ In addition, the MOH engaged partner laboratories, including the CDC-supported laboratories at the Kenya Medical Research Institute (KEMRI), to support laboratory confirmation of suspect cases.

Mpox testing in the Democratic Republic of Congo (DRC), the epicenter of the current clade I mpox outbreak, has demonstrated a high proportion of negative results, indicating alternative diagnoses in some persons with suspected mpox. The differential diagnosis for mpox, which causes a rash illness, is broad, but primary among the conditions that confound the clinical diagnosis is varicella (chickenpox) caused by varicella zoster virus (VZV). Varicella is a common rash illness in children and young adults that causes full body pox-like skin lesions and can be difficult to discern from a mpox rash.^2^ In DRC, confirmed varicella cases have been identified through mpox surveillance.^3^

Childhood varicella vaccination is not routinely done in Kenya, resulting in a large susceptible population.^4^ However, little has been published about varicella in Kenya,^5,6^ although routine surveillance reports suggest a seasonal pattern with a peak in September to November. To support the MOH’s mpox response, the CDC-supported KEMRI laboratories also conducted VZV PCR test, given the clinical overlap and high mpox test negativity rate early on among suspected cases.

## The Study

Specimens for this analysis were collected as part of the MOH’s mpox outbreak response in Kenya. A suspected mpox case was an unexplained rash in a person with at least one additional constitutional symptom and recent travel to another country experiencing a mpox outbreak, or contact with such a person, in the past three weeks.^1^ A probable case was a suspected case with contact with a probable or confirmed case or multiple or casual sexual partners in the past three weeks, or a positive orthopoxvirus test that was non-specific for monkeypox virus (MPXV). A confirmed case was a person with a positive MPXV PCR or genomic sequencing result.

Skin swabs were collected from lesions on ≥2 locations on a patient’s body using a synthetic swab that were placed together in the same media per patient and transported to the National Virology Reference Laboratory (NVRL) in Nairobi or the CDC-supported KEMRI laboratory in Kisumu. Samples sent to the NVRL were aliquoted and sent to the CDC-supported KEMRI laboratory in Nairobi for confirmation testing. Basic demographic information on patients was shared with the specimen aliquot.

MPXV PCR testing was performed using the CDC-developed assay^7^ and VZV testing was performed using primers targeting the VZV ORF-29 gene.^8^ Additionally, genomic sequencing of VZV-positive specimens was done using a targeted probe capture virus genome sequencing method, VirCapSeq-VERT^9^ and the Twist Bioscience NGS library prep (Twist Bioscience, CA) in a NextSeq2000 (Illumina Inc). FastQ files were analyzed online in CZID,^10^ and run files aligned using MAFFTv7 and visually inspected using BioEdit v7.2.5.^11^ Neighbor joining phylogenetic tree was inferred in SplitsTree v 6.4.7,^12^ with Kenya sequences alongside other genomes downloaded from GenBank-virus (https://www.ncbi.nlm.nih.gov/labs/virus/vssi/#/).

Between July and December 11, 2024, a total of 325 patients with suspected cases of mpox provided swabs for testing, and of these, 277 (85%) were tested at the CDC-supported KEMRI laboratories and had VZV testing. A total of 170 (61%) were positive for VZV, including four co-infections with mpox (Figure 1a). VZV percent positivity ranged from 34% among children ≤4 years to 69% among persons aged 20-49 years. The median age of patients with varicella was 23 years (interquartile range: 15-33 years), and 115 (68%) were males (Table 1). Patients with varicella were recorded in 32 (89%) of 36 counties reporting suspected mpox cases, with Nairobi County reporting the highest numbers (58; 34%) (Figure 1b). Although data on clinical outcomes was limited, one death occurred in a teenage female from Western Kenya who had a fever, full-body pustular rashes, and had tested VZV positive.

**Table 1.** Basic demographic characteristics of patients positive for varicella zoster virus (VZV) and monkeypox virus (MPXV) – Kenya, July-December 2024 (N = 277)

| Characteristic | VZV*<br>n = 170 | MPXV*<br>n = 26<sup>†</sup> |
| --- | --- | --- |
| Age Group |  |  |
| ≤4 years | 12 (7%) | 1 (4%) |
| 5-19 years | 50 (29%) | 4 (16%) |
| 20-49 years | 98 (58%) | 20 (80%) |
| ≥50 years | 10 (6%) | 0 (0%) |
| Missing | 0 | 1 |
| Sex |  |  |
| Female | 54 (32%) | 16 (62%) |
| Male | 115 (68%) | 10 (38%) |
| Missing | 1 | 0 |

\* Four patients were positive for VZV and MPXV
<sup>†</sup> During July to December 11, 2024, there were 28 total confirmed mpox cases in Kenya, of which 26 were confirmed at the CDC-supported Kenya Medical Research Institute laboratories.

**Figure 1.**
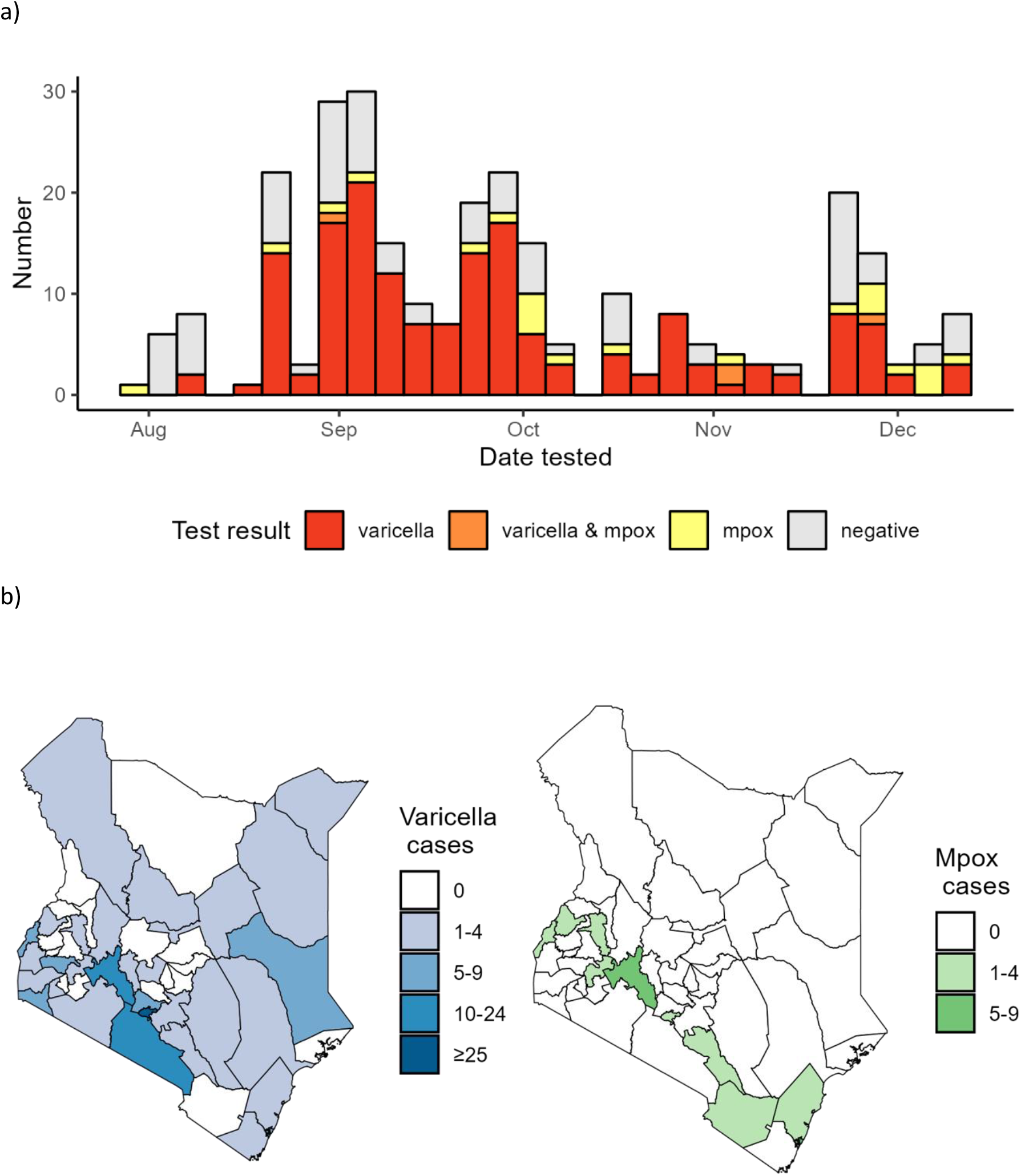
Timing (a) and geographic distribution (b) of patients testing positive for varicella zoster virus and monkeypox virus – Kenya, July-December 2024

Genomic sequencing was performed on 10/170 VZV-positive specimens. Completed genomes with coverage >85% were generated from eight of the ten (80%) specimens, all of which aligned with clade 5 human alphaherpesvirus 3 (aka VZV) (Figure 2). The Kenya sequences clustered alongside other sequences from sub-Saharan Africa as well as other regions of the world.

**Figure 2.**
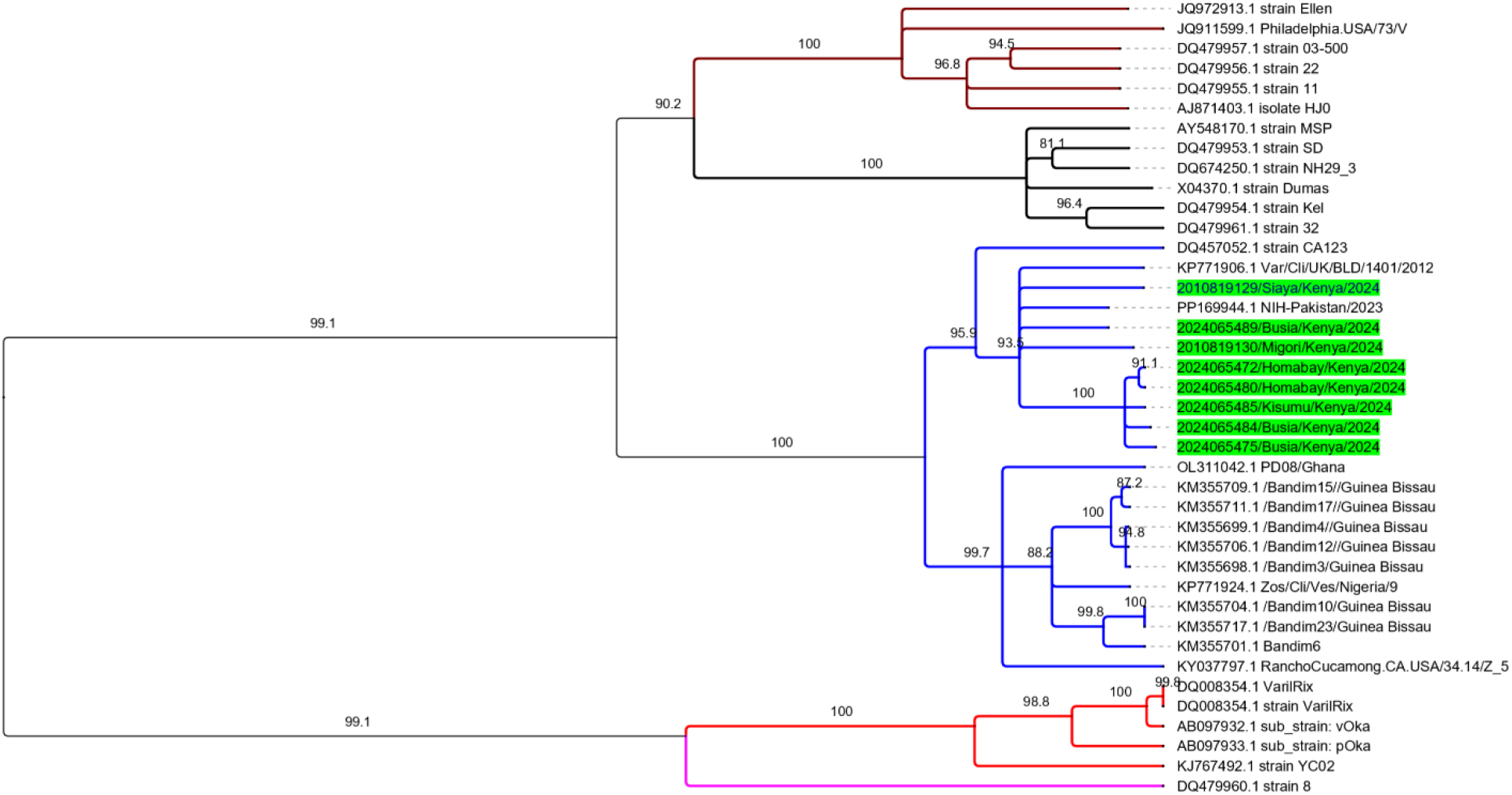
UPGMA phylogenetic tree showing Kenya varicella zoster virus (VZV) sequences alongside other sequences representing the 5 main VZV clades. Branches colored black represent genomes from clade 1, red represent clade 2, maroon represent clade 3, magenta represent clade 4 and blue represent clade 5. Kenya sequences are highlighted in lime color on the phylogenetic tree. Figures at the node of the branches are percent bootstrap values computed at 1,000 replicates.

## Conclusions

Testing suspected mpox cases for VZV during a mpox outbreak in Kenya improved disease intelligence, highlighting the importance of enhanced laboratory capacity to test for various pathogens during outbreak responses. Supporting this capacity in other countries experiencing mpox outbreaks could similarly reduce the proportion of unknown diagnoses in those who tested mpox negative. The high prevalence of varicella among patients with suspected mpox cases demonstrates the difficulty of untangling these diseases based on clinical presentation alone. Further exploration of clinical and epidemiological symptom differences could be helpful in defining the mpox outbreak case definition in Kenya. Co-infection with VZV and MPXV was also seen, which has also been observed elsewhere.^13^

These findings also highlighted a potential substantial burden of varicella in Kenya. Evidence from Africa suggests that varicella is ubiquitous by adulthood. Unlike in countries in temperate climates where varicella is a childhood disease, varicella might be most common in early adulthood in Kenya based on these findings.^14^ While severe disease and mortality from varicella is lower than other common vaccine preventable infections (e.g., measles), complications are more common among persons infected with HIV, which is a major public health problem in Kenya. Moreover, varicella can be teratogenic and causes more severe illness in adults. During this investigation, at least one death occurred in a varicella case. Dedicated studies that better characterize the public health burden of varicella in Kenya could inform discussions on varicella vaccination in Kenya.^15^

The Kenya genomes clustered with viruses from clade 5 that have been reported in parts of sub-Saharan Africa. Although the number of analyzed sequences was small, there was no unique clustering pattern by county to suggest a unique introduction of a new virus strain in Kenya. Despite this limitation, given the paucity of data from Eastern Africa on VZV, this additional genomic information could help bridge the gap.

As with many studies conducted during public health emergency responses, data completeness was a limitation because staff responsible for data collection were also responding to other urgent demands of the response. Hence, additional clinical features that could help characterize rashes of varicella and mpox were not available. Additionally, this study among patients with suspect mpox cases cannot provide information about the burden of varicella in the general population in Kenya.

The findings from this project demonstrate the importance of enhancing laboratory capacities to improve disease detection and response. Kenya has expanded the laboratory capacity of its public health reference laboratories, including that of the NVRL, through the addition of molecular testing and collaborations with partner laboratories such as the CDC-supported laboratories. Furthermore, CDC supports health security through laboratory network strengthening across the country to improve the ability to rapidly confirm outbreaks in Kenya. The addition of testing for other common rash illnesses like measles could further help define the proportion of suspect mpox cases caused by other causes.

## Data Availability

All data produced in the present work are contained in the manuscript

## CDC funding statement

This project was financially supported by the Centers for Disease Control and Prevention (CDC) under the terms of a cooperative agreement with the Association for Public Health Laboratories (APHL) (CoAg number: GH000032).

## Authorship disclaimer

The findings and conclusions in this report are those of the authors and do not necessarily represent the official position of the funding agencies.

## Acknowledgements

We thank the Kenya Ministry of Health staff for their work in responding to the mpox outbreak, Diagnostic and Laboratory Systems Program staff at the CDC-supported KEMRI laboratories in supporting the testing during this outbreak. We also thank Dr. Kenneth Wickiser and Jack Collins of the Global Alliance for Preventing Pandemics (GAPP) as well as other staff who supported the training on use of VirCapSeq-VERT for viral sequencing.

